# Molecular profiling of neuronal extracellular vesicles reveals brain tissue specific signals

**DOI:** 10.1101/2025.01.23.25320909

**Authors:** Vrinda Kalia, Gabriela Jackson, Regina J. Dominguez, Brismar Pinto-Pacheco, Tessa Bloomquist, Julia Furnari, Matei Banu, Olga Volpert, Katherine E. Manz, Douglas I. Walker, Kurt D. Pennell, Peter D. Canoll, Jeffrey N. Bruce, Erez Eitan, Haotian Wu, Andrea A. Baccarelli

## Abstract

Extracellular vesicles (EVs) released by neurons (nEVs) provide an opportunity to measure biomarkers from the brain circulating in the periphery. No study yet has directly compared molecular cargo in brain tissue to nEVs found in circulation in humans. We compared the levels microRNAs and environmental chemicals because microRNAs are one of the most studied nEV cargoes and offer great potential as biomarkers and environmental chemical load in nEVs is understudied and could reveal levels of chemicals in the brain. To do so, we leveraged matched sets of brain tissue and serum, and isolated serum total EVs and serum nEVs. We also generated and compared metabolomic profiles in a different set of matched serum, serum total EVs, and serum nEVs since metabolite cargo in nEVs is also understudied but could offer potential biomarkers. Highly expressed brain tissue miRNAs showed stronger correlations with nEVs than serum or total EVs. We detected several environmental chemical pollutant classes in nEVs. The chemical pollutant concentrations in nEVs were more strongly correlated with brain tissue levels than those observed between brain tissue and serum or total EVs. We also detected several endogenous metabolite classes in nEVs. Compared to serum and total EVs, there was enrichment of metabolites with known signaling roles, such as bile acids, oleic acid, phosphatidylserine, and isoprenoids. We provide evidence that nEV cargo is closely correlated to brain tissue content, further supporting their utility as a brain liquid biopsy.

## Introduction

Extracellular vesicles (EVs) are a heterogenous group of membrane bound vesicles that play an important role in cell-to-cell communication^1^. They can carry a variety of molecules - nucleic acids, lipids, metabolites, and proteins - which can affect cellular function in a recipient cell^1,2^. EVs released by neurons (nEVs), have been shown to influence cell function in regions of the brain distant from their point of origin^3^, propagate tau pathology^4,5^, and even regulate activation of glial cells^6^. NEVs can cross the blood-brain-barrier in both directions allowing for drugs to be delivered to the brain^7–9^ and for signals from the brain to reach the periphery^10–12^, providing a means for communication between the brain and the rest of the body. By enriching for nEVs in the periphery and analyzing their cargo, researchers have proposed nEV cargo as biomarkers of several neurodegenerative diseases^13^, including Alzheimer’s disease^14,15^, Parkinson’s disease^16,17^, and frontotemporal dementia^18^. Peripheral nEV cargo has also been linked to effects of modifiable factors, like exercise, in Alzheimer’s disease^19^. Some other applications of nEVs in observational studies have identified abnormal iron metabolism in restless leg syndrome^20^, dysregulated brain-gut-axis in people with IBS^21^, abnormal mitochondrial proteins in nEVs from people with major depressive disorder^22^, and predicted cognitive decline in people with HIV^23^. In a mouse study, researchers reported moderate to strong positive correlation between nEV pathological protein cargo and brain pathological burden^24^. Together, there is ample evidence that nEVs, and their cargo, provide an opportunity to discover biomarkers of neurological disease and offer insight into brain-periphery communications; however, no study yet has compared the cargo in circulating nEVs to their profiles in brain tissue in humans.

Similar to other EVs, nEVs harbor several classes of molecules^25^. MicroRNAs (miRNAs) are a class of noncoding RNA that regulate gene expression by targeting messenger RNA (mRNA) to induce degradation or by blocking translation^26^. They can be secreted by a cell into extracellular fluids, encapsulated in EVs or bound to proteins, and alter gene expression in a recipient cell, thus, acting as chemical messengers, mediating cell-to-cell communication^27^. Several studies have also measured nEV protein cargo^15,28^ and nEV lipid cargo^29^ as potential biomarkers of disease. While some studies have explored metabolites as cargo in CNS derived EVs^30^, no study has explored them in circulating nEVs.

Environmental chemical exposures have been linked to several neurological outcomes and diseases. Exposure to known pollutants like pesticides, fungicides, insecticides, air pollutants, and organic solvents have been associated with neurodegeneration^31^ and poor neurodevelopmental outcomes^32^. Most observational studies linking environmental chemical exposure to neurological health have measured chemicals in easily accessible compartments such as plasma, serum, urine, hair, and nails. While these compartments provide an aggregate measure of exposure, they are not specific to brain tissue. The mechanisms through which chemicals can enter the brain is highly regulated, particularly by the tight junctions between epithelial and endothelial cells at the blood-brain barrier and the blood-CSF barrier^33–35^. Thus, measuring these chemicals in nEVs could offer a better opportunity to capture exposure levels in the target organ of interest, the brain. Yet, no published study has described environmental chemicals in nEVs.

In this study, we leverage matched brain tissue and serum samples to measure miRNAs and environmental chemicals in four compartments: brain tissue, nEVs, total EVs, and serum (Figure 1). We then determined how levels of miRNA and environmental chemicals in brain tissue correlated with those in nEVs and compare this relationship to that between brain tissue and total EVs or serum. Further, we compared untargeted metabolomic profiles in nEVs, total EVs, and serum, from matched serum samples, since we did not have enough matched brain tissue and serum. Together, results from this qualitative analysis show the utility of nEVs as a window into brain health and provide evidence of their utility in measuring the burden of environmental chemicals in the brain.

**Figure 1.**
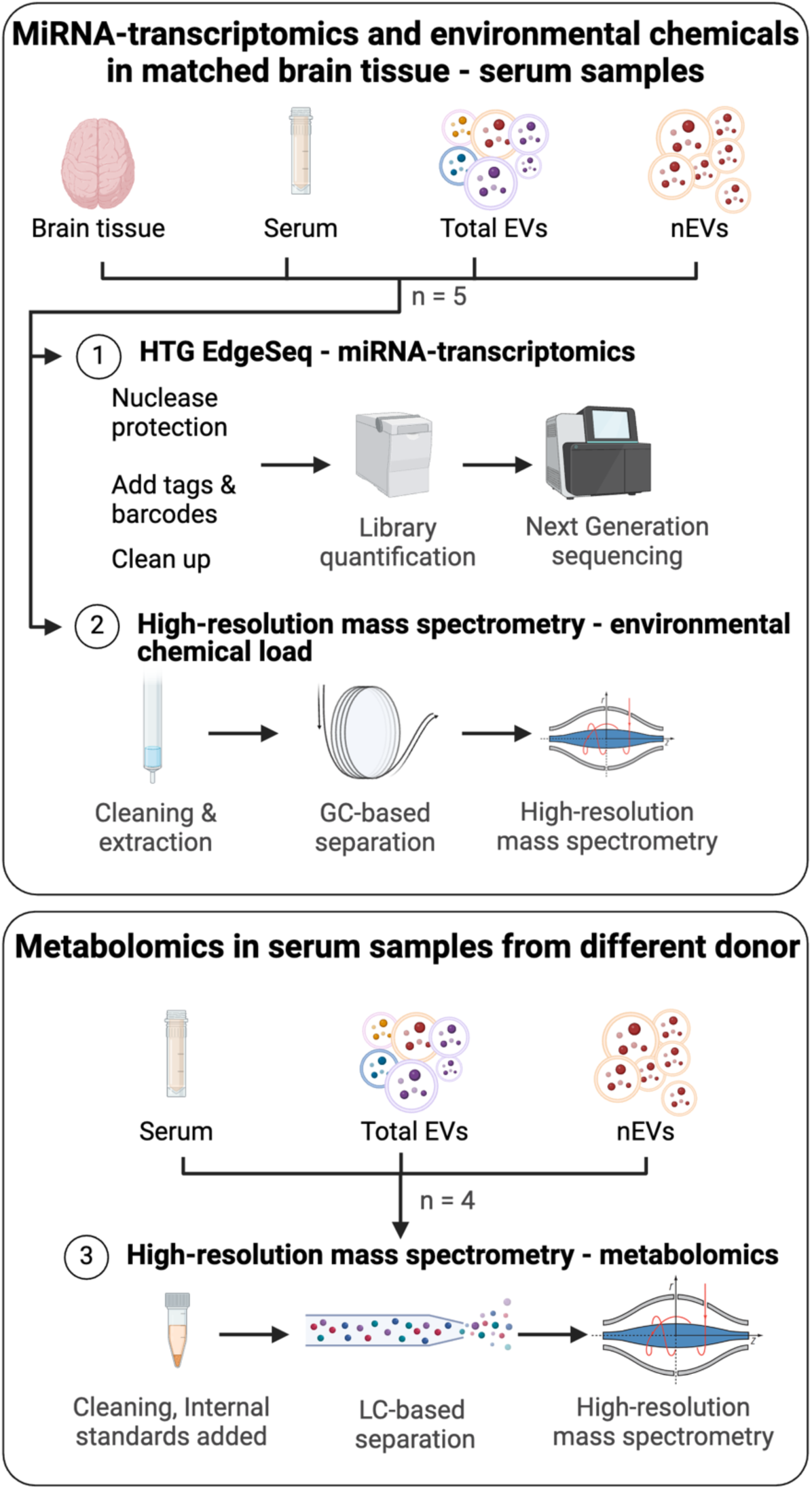
Study design. MiRNA: micro RNAs, EVs: extracellular vesicles, nEVs: neuronal EVs, GC: gas chromatography, LC: liquid chromatography.

## Methods

### 1. Samples

Five de-identified matched brain tissue and serum samples were obtained from the Bartoli Brain Tumor Laboratory under a protocol approved by the IRB at Columbia University Irving Medical Center. The samples were collected intraoperatively from either epilepsy or glioma brain tumor patients and stored in liquid nitrogen until needed for processing (S Table 1). Convenient unfiltered serum sample from an HIV-negative, male donor, in his 70’s, was obtained from BioIVT (Hicksville, NY). The sample tested negative for several infections tested.

### 2. Isolation of total EVs

Total EVs were isolated from 500uL of serum using the ExoQuick Ultra kit (System Biosciences, Palo Alto, CA) following the manufacturer’s recommendation. Briefly, samples were centrifuged at 3,000g for 15 minutes to remove cellular debris and 134 µL of ExoQuick precipitation solution was added to the supernatant. The mixture was inverted a few times and allowed to incubate on ice for 30 minutes and then centrifuged at 3000g for 10 minutes at 4°C. After aspirating the supernatant, the EVs in the pellet were resuspended using 200µL of buffer A. The purification column was pre-washed with buffer B, and then the sample along with 100 µL of buffer B were loaded onto the column. The sample on the column was mixed on a rotating shaker for 5 minutes, placed in a collection tube, and then centrifuged at 1000 g for 30 seconds to collect the flow through containing purified EVs.

### 3. Isolation of nEVs

NEVs were isolated from 500 µL of serum using the ExoSORT isolation kit (NeuroDex, Natick, MA)^15,36^, following manufacturer’s protocol. Briefly, total EVs were precipitated using NeuroDex Total EV isolation reagent, reconstituted in binding buffer and incubated overnight at 4°C with magnetic beads decorated with neuron-specific antibodies^15,36^. After nEV capture, the beads were washed three times on a magnetic separator with NeuroDex Wash Buffer. Beads, with captured nEVs, were air-dried and either stored for exposomics and metabolomic analyses or removed from beads using NeuroDex Elution buffer followed by lysis and miRNA extraction for sequencing.

### 4. nEV tetraspanins and relevant proteins

The tetraspanin profile of nEVs and total EVs was performed using a modified Luminex assay to determine levels of pan-EV tetraspanins CD63, CD9, and CD81^37^. Levels of neuronal proteins synaptophysin (SYP) and Repulsive Guidance Molecule a (RGMa), and a common serum contaminant, human serum albumin (HSA), were measured using enzyme linked immune sorbent assays (ELISA) kits (R&D Systems DuoSet ELISA, cat. No. DY1455, and DY245905) following manufacturer guidelines.

### 5. nEV concentration and size distribution measurements

The concentration and size distribution of nEVs and total EVs were determined using the ViewSizer 3000 (Horiba, MA) using methods previously described^38^. All samples were diluted at 1:1000 with 3 kDa filtered PBS and loaded into a cuvette for analysis. Twenty-five videos (30 frames per second, 300 frames per video) were recorded at 22°C with the following recording parameters: blue laser (445 nm), 210 mW; green laser (520 nm), 12 mW; red laser (635 nm), 8 mW; exposure,15 ms; camera gain, 30 dB. Samples were processed with the Main Chart in “LogBinSilica” and integrated in the range [50, 1000] nm. The summary of size distribution and particle concentration were extracted. Two technical replicates were run, and their average was reported.

### 6. nEV imaging

Transmission electron microscopy was used to visualize nEVs and total EVs following methods previously described^39^. Briefly, 5uL of EV suspension was placed on a formvar/carbon-coated grid and allowed to settle for 60 seconds. The samples were negative stained and blotted using 1.5% uranyl acetate in water and allowed to air dry at room temperature. Grids were imaged with a JEOL JSM 1400 (JEOL, USA, Ltd, Peabody, MA) transmission electron microscope, operating at 100 kV, and images were captured on a Veleta 2K × 2K CCD camera (Olympus-SIS, Munich, Germany). 10-15 representative images were captured of three randomly selected grid areas per sample at 50,000 and 100,000× lens magnification. Scale bars reflect the camera magnification.

### 7. miRNA sequencing

MiRNAs were sequenced using the HTG EdgeSeq miRNA Whole Transcriptome Assay, which quantifies the expression of 2,083 human miRNAs (HTG Molecular Diagnostics, Inc., Tucson, AZ, USA). Brain tissue (5 mg), serum (30 uL), total EVs (30 µL), and nEVs (30 µL), were mixed with 30 µL of HTG biospecimen lysis buffer. The brain tissue was then processed in a bead beater to disrupt the tissue, and the supernatant was used for analysis. Three samples were included as internal technical replicates for quality control purposes.

### 8. Gas chromatography coupled high-resolution mass spectrometry (environmental chemical analysis)

Brain tissue (weighing between 40 and 170 mg), 300 uL of serum, 300 uL of total EVs in buffer and nEVs on beads were analyzed for levels of 120 chemicals using gas chromatography coupled high-resolution mass spectrometry. Brain tissue was processed in a bead beater with acetonitrile to extract the chemicals. Briefly, yttria stabilized zirconium oxide beads (∼10 beads, 0.5 mm diameter, Next Advance, Troy, NY) were added to the tissue and samples were placed in a bead beater (Next Advance Bullet Blender Storm, Troy, NY) set at speed 6 m/s for 5 minutes. Serum and total EVs, and nEVs were extracted using a QuEChERS extraction^40^. Briefly, samples were sonicated in 1 mL hexane:acetone:dichloromethane and transferred to a 2 mL QuEChERS containing 150 mg MgSO4 and 50 mg C18. The supernatant was transferred to a new vial and the QuEChERS tube was washed with 0.5 mL hexane:acetone:dichloromethane twice. The extract was evaporated under nitrogen to a final volume of 200-300 µl hexane using an Organomation 30 position Multivap Nitrogen Evaporator (Organomation Associates Inc.) and transferred to a glass autosampler vial. Sample extracts were analyzed using a high-resolution Thermo Q Exactive Orbitrap MS equipped with a Thermo Trace 1300 GC and a TriPlus RSH Autosampler (details are provided in S Table 2).

### 9. Liquid chromatography coupled high-resolution mass spectrometry (metabolomics)

Serum (100uL), total EVs (in 150 uL buffer), and nEVs (on beads) were used to generate metabolomic profiles using liquid chromatography with high-resolution mass spectrometry. The method has been previously published^41^. We included buffer B as a blank to filter non-sample signal from total EVs. In brief, the samples were extracted using acetonitrile with internal standards. The extract was analyzed using two different analytical columns, HILIC and C18, each operated under positive and negative electrospray ionization (four assays total). An acetonitrile gradient with formic acid or 10 mM ammonium acetate adjusted to pH of 9.5 was used for HILIC positive and negative, respectively, an acetonitrile gradient with formic acid or 10 mM ammonium acetate was used for C18 positive and negative, respectively. For each analysis, 10 µL of the sample extract was injected on each column. Mass spectral data were generated on a Thermo Scientific HFX orbitrap mass spectrometer in full scan mode, scanning for mass range 85 to 1250 Da operated at a resolution of 120,000. All raw mass spectral data were extracted using the R packages apLCMS^42^ and xMSanalyzer^43^. Through the analysis pipeline, four feature tables were generated: HILIC (+ ESI), HILIC (- ESI), C18 (+ ESI), and C18 (- ESI).

### 10. Data analysis

All analyses were performed using R (version 3.6.3) unless stated otherwise.

#### 10.1. MiRNA data normalization

MiRNAs detected with a raw count greater than 30 in more than 30% of the samples were retained for analysis to remove miRNA with low abundance. Out of the 2083 miRNA measured, 1840 were retained. We used trimmed mean of M-value (TMM) method in the edgeR (version 3.40.2)^44^ package to normalize read counts and calculate normalized counts-per-million for each miRNA.

#### 10.2. Exposomics data processing

The detection rates of chemicals in each compartment (brain tissue, serum, nEVs, total EVs) were determined and chemicals detected in all compartments for at least four individuals were retained for downstream analysis (18 chemicals). Values below the limit of detection in the filtered dataset were replaced with half of the value of either the limit of detection or lowest detection, whichever was smaller.

#### 10.3. MiRNA pathway analysis

We used DIANA miRpath (version 4.0)^45^ to find target genes for the top 100 most abundant miRNA in each of the compartments. Target genes for the top 100 most expressed miRNA were determined using the TarBase v8 database, a collection of experimentally supported miRNA– gene interactions. A miRNA-centric analysis was performed using the following setting: 1) long non-coding RNA were included among targets, 2) we used miRbase-v22.1 as annotation source, and Kyoto Encyclopedia of Genes and Genomes (KEGG) as pathways database, 3) merging methods was set to “genes union” and testing method to “classic analysis”, 4) we applied a false discovery rate (FDR) correction with p-value threshold set to < 0.05. Of the 100 miRNAs tested in each compartment, we did not find targets for 9 miRNAs in brain tissue, 46 miRNAs in total EVs, 57 miRNAs in nEVs, and 58 miRNAs in serum. Therefore, to determine the importance of a target pathway in each compartment, while accounting for the variability in the number of miRNAs with known targets, we calculated the ratio of the number of miRNAs that target a pathway and the total number of miRNAs that had a known target within each compartment. We then compared these ratios for pathways across the four compartments using an ANOVA and Tukey post hoc test. We also tested for pathways targeted by miRNAs that were correlated between nEVs and brain tissue (absolute correlation ≥0.8) using the same settings.

#### 10.4. Correlation between molecules in brain tissue and other compartments

We determined the correlation between the molecules in brain tissue (top 30 miRNAs, based on mean level in tissue, and environmental chemicals), and their levels in the three other compartments using Spearman’s rank correlation. Further, we investigated whether certain physicochemical properties were related to the correlation of chemicals between brain tissue and the other compartments. We used the predicted octanol-water partition coefficient (logP), predicted biodegradation half-life, average mass, and density for chemicals, obtained from the EPA CompTox dashboard,^46^ and determined the correlation between these properties and computed correlation between each compartment and brain tissue using Spearman’s rank correlation.

#### 10.5. Metabolomics data processing and annotation

Data from the four chromatographic columns and ionization modes were handled similarly. Blank filtration was performed to remove signal from buffers used to collect EVs. The threshold for filtration was determined as 2 × (*Blank_mean_* + (3 × *Blank_standard deviation_*)) for each feature detected^47^. The filtered features were used for downstream analysis after replacing zero values, considered below the limit of detection, with half the lowest intensity detected for that feature. The intensities were log2 transformed for statistical analysis. The *prcomp()* function was used to conduct principal component analysis and biplots were created using the factoextra package (version 1.0.7) to look at patterns in data based on the sample type. Metabolic features were annotated using in-house reference libraries first. Features that remained unannotated were assigned annotations using xMSannotator^48^ with the human metabolome database as the reference. Only features with an annotation confidence score greater 1 were retained for downstream analysis. Metabolite annotation confidence was assigned based on the Schymanski scale^49^ where level 1 corresponds to a confirmed structure identified through MS/MS and/or comparison to an authentic standard; level 2 to a probable structure identified through spectral matches to a database; level 3 to a putative identification with a speculative structure; level 4 to an unequivocal molecular formula but with insufficient evidence to propose a structure; and level 5 to an exact mass but not enough information to assign a formula. The metabolomic data comprised 17,889 features from the HILIC (+ ESI) column, 11,965 features from the HILIC (-ESI) column, 17,392 features from the C18 (+ ESI), and 9,023 features from the C18 (-ESI) column. After blank filtration, 6,956 (39%) features were retained from the HILIC (+ESI) column, 2,280 features (20%) from the HILIC (-ESI) column, 6,257 features (36%) from the C18 (+ESI) column, and 2,366 features (25%) from the C18 (-ESI) column. Of the features remaining after blank filtration, 3,507 features (20%) were assigned an annotation, 248 (1.5%) at level 1 and 1,673 (10%) at level 3. A biplot of PC1 and PC2 from a principal component analysis showed clustering based on sample type (S Figure 3).

#### 10.6. Metabolite class membership

Metabolite class membership was determined using RefMet, hosted on the metabolomics workbench website^50^. The super class, main class, and sub class membership was obtained using annotated metabolite names.

#### 10.7. Enrichment Analysis

The main class enrichment of the top 10% of metabolites was determined using the “enrichment analysis” module in MetaboAnalyst (version 6.0)^51^. An FDR of 20% was used to filter noteworthy enrichment and FDR at 5% was considered significant.

#### 10.8. Analysis of differential levels of super classes and metabolites

Multiple ANOVAs followed by a post hoc Tukey test were used to find metabolite super classes that were differentially abundant in the three compartments. The sub classes of the metabolites that were significantly higher in nEV compared to serum and total EVs were determined.

## Results

### Characterization of nEVs and total EVs

Compared to total EVs, nEVs had detectable, but lower levels of tetraspanins CD9, CD63, and CD81 and higher levels of neuronal proteins SYP and RGMa (S Figure 1 A & B). nEVs had fewer particles (number of EVs) than total EVs and showed a similar size distribution (S Figure 1C) although the median particle size was larger (nEV median = 173.48 nm and total EV = 114.76 nm, S Figure 1D). Visual inspection of electron micrographs showed cup shaped vesicles in both total EVs and nEVs isolated (S Figure 1E).

### NEVs miRNAs are correlated with brain tissue miRNAs and target similar pathways

The top 30 statistically significant pathways, enriched by the top 100 most expressed miRNA targets in brain tissue, included several neurologically relevant pathways: pathways of neurodegeneration (multiple diseases), amyotrophic lateral sclerosis (ALS), Huntington’s disease (HD), Parkinson’s disease (PD), Alzheimer’s disease (AD), axon guidance, spinocerebellar ataxia, and neurotrophin signaling pathway (Figure 2A, S Table 3). Among the enriched pathways, the mean proportion of miRNAs that target each pathway in the brain was most closely related with nEVs, followed by total EVs, and then serum (S Table 3).

**Figure 2.**
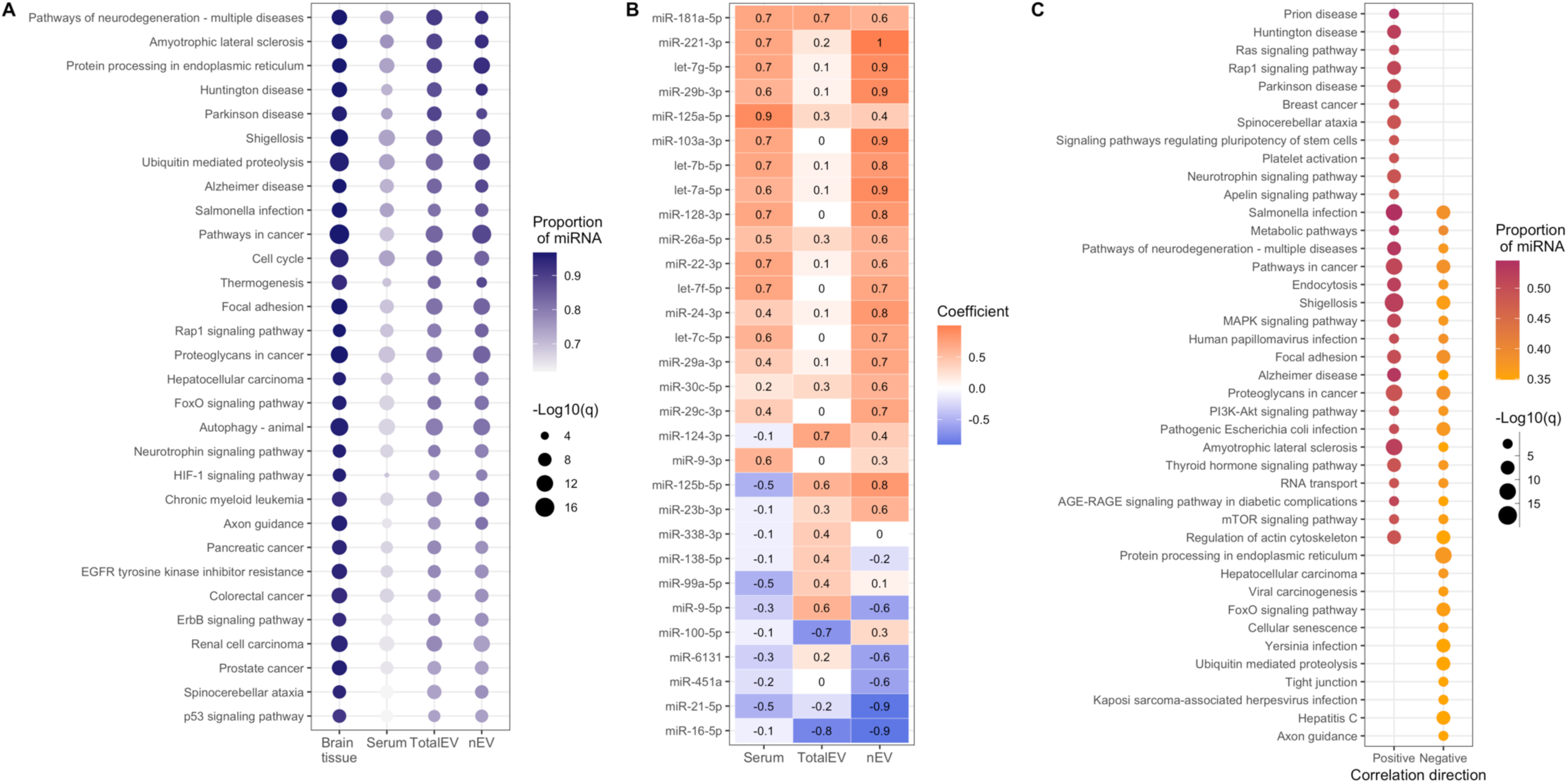
Brain tissue miRNA profiles compared to profiles in serum, total EVs, and nEVs. In A, the top 30 pathways (based on FDR q-value) targeted by the top 100 most abundant miRNA measured in brain tissue. The intensity of the color represents the proportion of top 100 miRNA in each compartment that target the pathways and the size of the dot indicates the FDR q-value of statistical significance. In B, the correlation between the top 30 most abundant miRNA in brain tissue and their corresponding level in each compartment. The numbers and colors represent the correlation coefficient. In C, the top 30 pathways (based on number of miRNA) targeted by nEV miRNAs positively and negatively correlated with brain tissue (|rho| ≥ 0.8). The size and color represent similar attributes as in A.

The correlation between the top 30 most expressed miRNA in brain tissue and their expression in other compartments showed a greater number of positive correlations between brain tissue and nEVs compared to other two compartments (Figure 2B). The expression of miR-221-3p in the brain was perfectly correlated with its expression in nEVs (Spearman’s correlation coefficient (rs) = 1, p = 0.02), positively correlated with its expression in serum (rs = 0.7, p = 0.23) and not correlated with its expression in total EVs (rs = 0.2, p = 0.78). Other miRNA with strong positive correlation (rs ≥ 0.8) between brain tissue and nEVs included: let-7g-5p, miR-29b-3p, miR-103a-3p, let-7b-5p, let-7a-5p, miR-128-3p, miR-24-3p, and miR-125b-5p. The expression of miR-125a-5p in brain tissue was positively correlated with its expression in serum (rs = 0.9, p = 0.08) and modestly correlated with its expression in both nEVs (rs = 0.4, p = 0.52) and total EVs (rs = 0.3, p = 0.68). The correlation coefficients and p-values for all other miRNA analyzed can be found in S Table 4.

The miRNAs positively correlated between nEVs and brain tissue were enriched in several neurologically relevant pathways: pathways of neurodegeneration (multiple diseases), ALS, HD, PD, AD, axon guidance, spinocerebellar ataxia, and neurotrophin signaling pathway (Figure 2C). The miRNAs negatively correlated between nEVs and brain tissue were enriched in several neurologically relevant pathways: AD, pathways of neurodegeneration (multiple diseases), ALS, and axon guidance, some signaling pathways, and pathways related to viral infections (Figure 2C, S Table 5).

### Chemical load in brain tissue is correlated with their concentration in nEVs

Of the 121 chemicals tested, 12 were not detected in any sample, 60 were detected in at least one brain tissue sample tested, 64 were found in at least one nEV sample tested, 74 were found in at least one total EV sample tested, and 65 were found in at least one serum sample tested (S Table 7). Twenty-eight chemicals were found in all four compartments of at least one person and 18 were detected in all four compartments in at least four individuals (Figure 3A). Diethyl phthalate, DEET, and 2,3,7,8-tetrachlorodibenzofuran (2,3,7,8-TCDF) showed the highest absolute concentrations in the brain (S Table 8). Concentration of chemicals in brain tissue was consistently the lowest compared to the other three compartments (S Table 9).

**Figure 3.**
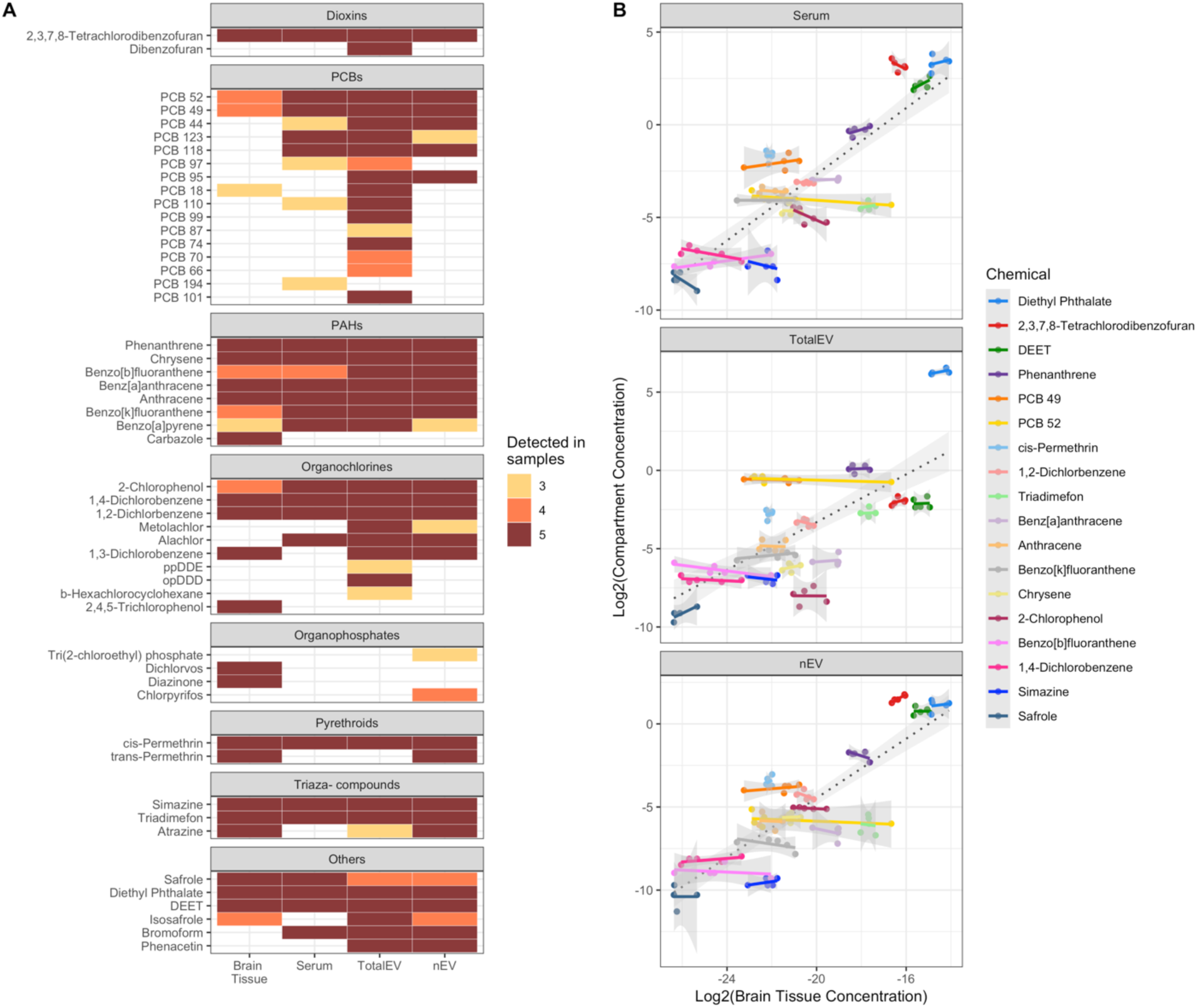
Environmental chemicals detected in each compartment and their correlation with levels in brain tissue. In A, the chemicals detected in at least 3 samples in any compartment. In B, for chemicals detected in at least 3 samples in all compartments, the relationship of their levels in brain tissue with serum, total EVs, and nEVs.

Looking at all chemicals, as a whole, the correlation between chemicals detected in the brain and their levels in the other three compartments was positive, with the strongest correlation between brain tissue and nEVs (rs = 0.72, p = 7.2e-16), followed by serum (rs = 0.7, p = 5.8e-15) and lowest for total EVs (rs = 0.58, p = 1.5e-09). For individual chemicals, the levels in brain tissue of the pollutants 2,3,7,8-TCDF (rs = 0.8), polychlorinated biphenyl 49 (PCB 49, rs = 0.8), and 1,4-dicholorobenzene (1,4-DCB, rs = 0.67) were positively correlated with their levels in nEVs. The levels of phenanthrene (rs = 0.8), benz[a]anthracene (rs = 0.7), and DEET (rs = 0.7) in the brain were positively correlated with their levels in serum. The correlation between brain tissue and total EVs for diethyl phthalate (rs = 0.8), 2,3,7,8-TCDF (rs = 0.7), and safrole (rs = 0.62) was the strongest positive relationship. None of the individual chemical correlations were statistically significant (S Table 10). The correlation of chemical concentrations between brain tissue and all compartments for all 18 chemicals can be found in S Table 10. Higher density chemicals were less correlated between serum and brain tissue (rs = −0.51, p = 0.03), but density did not impact the chemical correlations between brain and nEVs or total EVs. Chemical octanol-water partition coefficient, average mass, and biodegradation half-lives were not correlated with correlation coefficients from any of the brain-compartment correlations (S Table 11).

### Metabolomic profile of nEVs shows enrichment of metabolites with signaling roles

Of the annotated metabolites queried through RefMet, 888 (5%) were assigned a class membership. When comparing the super class members across compartments, 15 of the 17 classes were significantly different across all the compartments (Figure 4 A). Organic oxygen and organohalogen compounds were not significantly different across all compartments. Post hoc analysis showed that all super classes had highest levels in serum. When comparing total EVs and nEVs, 5 super classes were not significantly different, these were: organic oxygen compounds, organohalogen compounds, prenol lipids, sterol lipids, and sphingolipids. All other classes had higher levels in total EVs compared to nEVs (S table 12).

**Figure 4.**
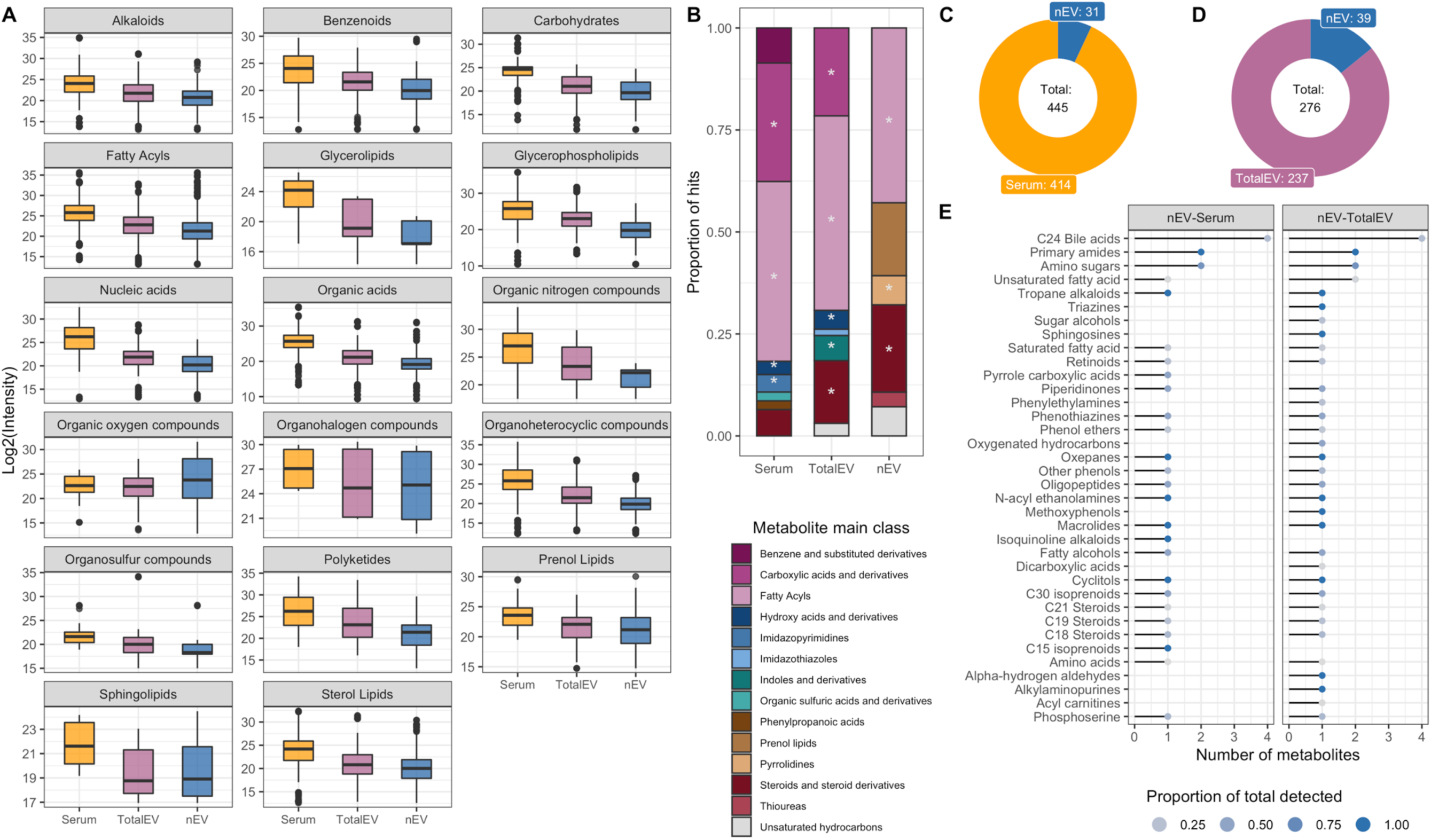
Metabolomic profiles of serum, total EVs, and nEVs. In A, the intensities of features within each super class of metabolites in each compartment. In B, main classes of metabolites represented by the top 10% of the most abundant metabolic features in each compartment with at least 2.5% of total metabolites tested. In C, the number of features with significantly different levels between nEVs and Serum, with each section of the donut indicating the number with higher intensity in the respective compartment, i.e., a total of 445 features were significantly different of which 31 were higher in nEV than serum. In D, the number of features with significantly different levels between nEVs and total EVs. In E, the sub classes of metabolic features with higher intensity in nEVs compared to serum and total EVs. The color of the dot represents the ratio of number metabolites higher in nEV over the total number of metabolites detected in each subclass.

Fatty acyls were the only main class enriched by the top 10% of most abundant metabolites in all three compartments (FDR<0.05). Carboxylic acid derivatives and hydroxy acids and derivatives were enriched in both the serum and total EV compartments while steroids and steroid derivatives were enriched in both the total EV and nEV compartments (FDR<0.05). Classes uniquely enriched to each compartment were imidazopyrimidines in serum, indoles and derivatives in total EVs, and pyrrolidines in nEVs. Other classes unique to each compartment but not statistically significant, included benzene and substituted derivatives, organic sulfuric acid and derivatives, and phenylpropanoic acids in serum; imidazothiozoles in total EVs; and prenol lipids, and thioureas in nEVs (Figure 4B, S table 13).

Testing for the difference in feature abundance between the three compartments revealed 445 metabolites significantly different between serum and nEVs, of which 31 had higher mean levels in nEVs (Figure 4C), and 276 significantly different metabolites between total EVs and nEVs, of which 39 had a higher mean in nEVs (Figure 4D). A total of 36 subclasses were represented by these metabolites with higher levels in nEVs (Figure 4E) and C24 bile acids had the highest number of members (n = 4) followed by primary amides (n = 2), amino sugars (n = 2) and unsaturated fatty acid (n = 2 when comparing nEVs and total EVs). The bile acids higher in both comparisons were: taurocholic acid (level 1), allocholic acid (level 1), and dehydrolithocholic acid (level 1). Chenodeoxycholic acid sulfate (level 3) was higher in nEVs compared to serum and beta-muricholic acid was higher in nEVs compared to total EVs. The primary amides were palmitamide (level 3) and oleamide (level 3); the amino sugars were glucosamine (level 3) and galactosamine (level 1); the unsaturated fatty acids were oleic acid (level 1, higher in both comparisons) and stearidonic acid (level 1, higher only between nEVs and EVs, S Figure 4, S table 14)

## Discussion

We present the first study of miRNA-transcriptomics, environmental chemical load, and metabolomics in nEVs, total EVs, and serum. By leveraging matched brain tissue and blood samples, we were able to compare these -omic profiles in the brain against the other three compartments. We found that the highly expressed brain tissue miRNAs were strongly correlated with nEV miRNAs and that the ranking of chemical concentrations in the brain tissue were preserved in nEVs. Thus, we provide evidence that for miRNA-transcriptomics, nEVs provide a better reflection of brain miRNA signal compared to serum or total EVs. We also report several metabolite classes in nEVs and enrichment of some bile acids, fatty acids, and isoprenoids in nEVs. The small sample size limited the statistical power in the analyses conducted; however, we were able to make qualitative assessments of the -omic profiles across the different compartments.

The most abundant miRNA detected in brain tissue targeted pathways related to neurological disease, neuronal function, and cancer. This was also observed for miRNAs abundant in nEVs and total EVs and, to a lesser extent, in serum. However, the correlation between the top 30 miRNAs in the brain and total EVs was weaker compared to the correlation between the brain and nEVs, suggesting that nEV miRNA profiles more accurately reflect brain tissue and are therefore better at informing gene changes associated with neurological diseases.

The nEV miRNA with the strongest positive correlation with the 30 most abundant brain miRNAs have previously been associated with several nervous-system-related outcomes, including Alzheimer’s disease (miR-29b-3p^52^, let-7b-5p^53^, miR-128-3p^54^), autism spectrum disorders (let-7g-5p, miR-103a-3p^55^, let-7b-5p^55^, miR-125b-5p^56^, let-7a-5p^57^), glioblastomas (miR-221-3p^58–60^, miR-128-3p^58^, miR-24-3p), pituitary adenomas (miR-128-3p^61^, miR-24-3p, let-7a-5p^61^), multiple sclerosis (let-7b-5p^62^), and onset of epilepsy (let-7b-5p^63^). Other miRNA (miR-103a-3p, miR-125a-5p^64^) also targeted brain-derived neurotrophic factor (BDNF), a key protein involved in the growth, development, and maintenance of neurons in the brain. All these miRNAs have been previously detected in extracellular space^65,66^, some of them encapsulated in EVs^67^ and exosomes^68,69^. Additionally, the pathways targeted by all nEV miRNAs positively correlated with brain tissue levels (rho ≥ 0.8) included several neurological diseases, cancers, and signaling pathways. The nEV miRNAs negatively correlated with brain tissue levels (rho ≤ −0.8) targeted fewer pathways related to neurological diseases but more pathways related to inflammation and infections. Thus, it is evident that the miRNAs detected in nEVs are relevant to neurological function and disease and have utility as biomarkers of neurological health disease.

We detected chemicals from several chemical classes, organochlorines, dioxins, PAHs, PCBs, pyrethroids and others, in all four compartments. While only a few chemicals in nEVs were positively correlated with their levels in brain tissue, there was an overall positive correlation between chemical levels, as a whole, in brain and nEVs suggesting that the relative levels of chemicals in the brain are reflected in their levels in nEVs. For example, based on concentration, the top three chemicals in brain tissue (diethyl phthalate, 2,3,7,8-TCDF, and DEET) were also the top three chemicals in nEVs. This hierarchy was also observed in the concentration of these chemicals in serum but not in total EVs.

Among the characteristics expected to influence the levels of chemicals detected in the brain are their lipophilicity, weight, half-life, and ability to cross the blood-brain barrier^70^. There was no clear relationship between any chemical physicochemical property tested and their correlation between brain tissue and nEV and total EVs. This may suggest that these physicochemical properties do not influence the levels of chemicals in EV released by neurons; however, our study might have been underpowered to find these relationships. The chemical density – mass per unit volume in g/cm^3^ – was negatively correlated with correlation coefficients between serum and brain tissue, i.e., molecules with larger density had a negative correlation between serum and brain tissue levels, suggesting better diffusion of less dense molecules into the brain^70^. We did not see a relationship between the correlation coefficient and lipophilicity or weight, which has been previously reported^70^. Some other factors that may influence the chemical concentrations and correlation include the time elapsed between exposure and blood draw, and underlying differences in the toxicokinetics that may influence the amount of chemical reaching the blood stream and its clearance from the body^71^.

The factors that may influence the concentration of chemical pollutants in nEVs have not yet been studied. The presence of chemicals maybe a result of packaging into EVs for removal from cells^72^. It may also be due to their presence in the cytosol, likely bound to proteins or lipid molecules, at the time of EV biogenesis^1^. It remains possible that circulating chemical molecules, especially, lipophilic molecules, may adhere to the surface of EVs in the periphery^73^. Since no other published study has looked at this relationship, future research should investigate the underlying mechanics of cellular packaging of chemicals into EVs.

Previous studies have reported a diversity of metabolites in total EVs. Studies have shown the presence of amino acids, tricarboxylic acid cycle intermediates, purines, fatty acids, ceramides, phospholipids, carnitines, and carbohydrates^74,75^. We also detected these metabolite classes in total EVs. Several metabolite classes were also detected in nEVs. Although the overall metabolite levels were lower in the nEVs compared to total EVs and serum, the diversity of detected metabolites suggests that nEVs offer a unique opportunity to assess the metabolic status of neurons and identify potential disease biomarkers. Among the top 10% of metabolites significantly enriched in nEVs, were fatty acyls, pyrrolidines, steroids and steroid derivatives.

The levels of several metabolites were higher in nEVs compared to serum and total EVs, including several membrane stabilizing molecules, steroids, bile acids, and other lipid molecules, suggesting enrichment of these metabolites in nEVs compared to serum and total EVs^76^. C-24 bile acids, such as taurocholic acid, allocholic acid, and chenodeoxycholic acid, play a role in bile-acid signaling pathways^77^ and their enrichment in nEVs may offer insight into the role of nEVs in the gut-brain-axis. Among metabolites enriched in nEVs was a phosphoserine, consistent with previous evidence that other phosphoserines were enriched in brain derived EVs compared to their tissue of origin^75^. Given their role in neurotransmission, signaling, and neuronal function, phosphoserines in nEVs could be potential biomarker candidates for neurological health and diseases^78^. A short chain C15 isoprenoid, farnesol, was also enriched in nEVs. Short-chain isoprenoids have been reported to play an important role in post-translational modification of proteins and several studies suggest targeting isoprenoid pathways for treatment of Alzheimer’s disease^79,80^. Its enrichment in nEVs may shed light into communication between the brain and periphery, given the effects of protein prenylation on cellular function^80^. We also saw enrichment of saturated fatty acid, lauric acid, and unsaturated fatty acids, oleic acid and stearidonic acid. Given that the dry weight of the adult brain is approximately 55% lipid, thirty five percent of which is made up by polyunsaturated fatty acids, the presence of fatty acids in nEVs is expected and previously documented in brain derived EVs^75^. Oleic acid, a monounsaturated fatty acid, is the primary fatty acid in the white matter of the brain^81^ and has been reported to play a role in nutrient sensing via dopaminergic neurons in the ventral tegmental area^82^. Its enrichment in peripheral nEVs indicates the presence of functionally important fatty acids which could offer mechanistic insight into neuronal function or disease. To the best of our knowledge, this is the first study to describe the metabolic cargo of nEVs, compared to serum and total EVs. Other studies are needed to corroborate our findings.

This is the first multiomic analysis of nEV miRNA, exposomic, and metabolomic cargo with matched comparisons. Despite the many novel findings from our analyses, our study suffered from a few notable weaknesses. First, our study had a limited sample size and was not powerful enough to detect small and moderate differences in molecules/chemicals between the compartments. Second, we did not have enough material to also conduct metabolomics in matched brain tissue and serum samples. So, while conveniently available serum was used to characterize the metabolic profile of nEVs, we were unable to deduce their relationship with the neuronal metabolome. Third, the brain tissue sections were usually taken from the cortex and did not represent the state of neurons across the whole brain. Fourth, we did not measure all potential environmental pollutants of concern, such as perfluorinated compounds. They have been detected in the brain and reported to be correlated with serum^83^. We also did not measure whether nanoplastics are detected in nEVs which also warrants investigation since they may be small enough^84^ to be packaged within EVs. However, these drawbacks do not take away from our findings and future studies should prioritize answering the unknown relationships identified through our study.

## Acknowledgements

V.K. was supported by NIH/NIA grant P20AG093975. H.W. and A.A.B. were supported by NIH/NIEHS grant R35ES031688.

## Data availability

Data available from corresponding author upon reasonable request.

