## Supplemental Figures for "Molecular profiling of neuronal extracellular vesicles reveals brain tissue specific signals"

Supplemental Figure 1. **Characterization of extracellular vesicles.** In A, we detected lower levels of CD9, CD63, and CD81 in nEVs compared to total EVs and the nEV depleted fraction. We found higher levels of neuronal specific proteins synaptophysin and RGMa compared to total EVs and the nEV depleted fraction. In B, we saw lower levels of albumin in nEVs and total EVs compared to the nEV depleted fraction. In C &D, there were lower number of particles in nEVs compared to nEV depleted fraction and total EVs. The distribution of particles across size was similar for total EVs and the nEV depleted fraction, while there were more larger particles in nEVs compared to the other two. In E, we visualized the nEVs and total EVs using transmission electron microscopy. SYP: synaptophysin, RGMa: repulsive guidance molecule a, D10: size below which 10% of particles was contained, D50: size below which 50% of particles was contained, D90: size below which 90% of particles was contained, MFI: mean fluorescence intensity, AU: arbitrary unit.

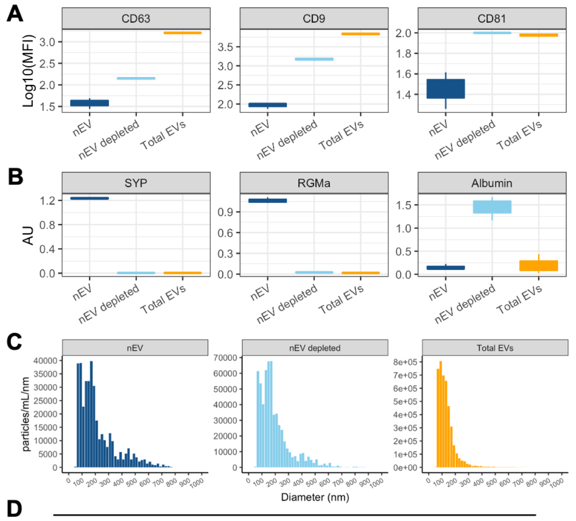


D

| Parameters | nEV | nEV depleted | Total EVs |
|---|---|---|---|
| D10 (nm) | 76.77 | 73.65 | 71.78 |
| D50 (nm) | 173.48 | 116.085 | 114.76 |
| D90 (nm) | 333.945 | 197.12 | 186.87 |
| Mode (nm) | 60.7 | 77.04 | 76.19 |
| Concentration (particles/mL) | 2.35E+09 | 1.57E+11 | 8.2E+10 |

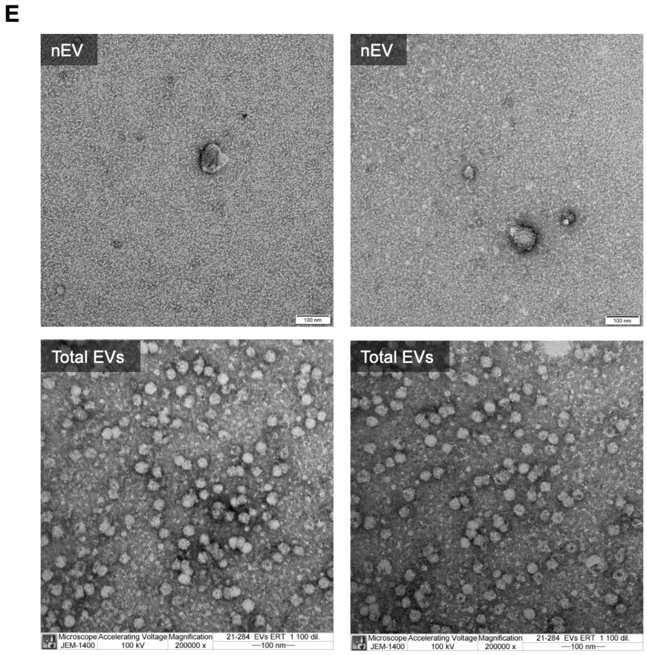

Supplemental Figure 2. **Correlation between brain tissue concentration and concentration in compartments.** The correlation coefficient from Spearman correlation analysis for each chemical level between brain tissue and compartment are shown. The red color indicates a positive association.

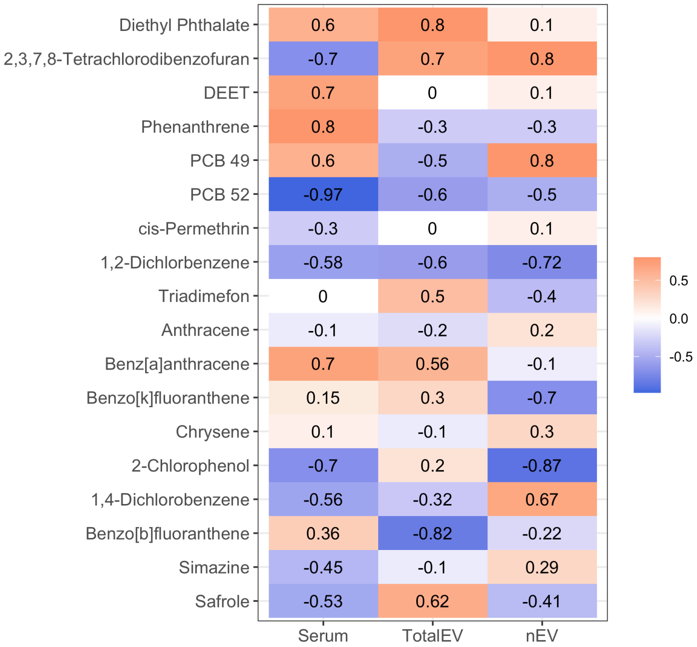

Supplemental Figure 3. Biplot of PC1 and PC2 generated from principal component analysis of metabolomics data shows clustering of samples based on compartment source. Data from HILIC (+ESI) and C18 (+ ESI) show better separation of clusters.

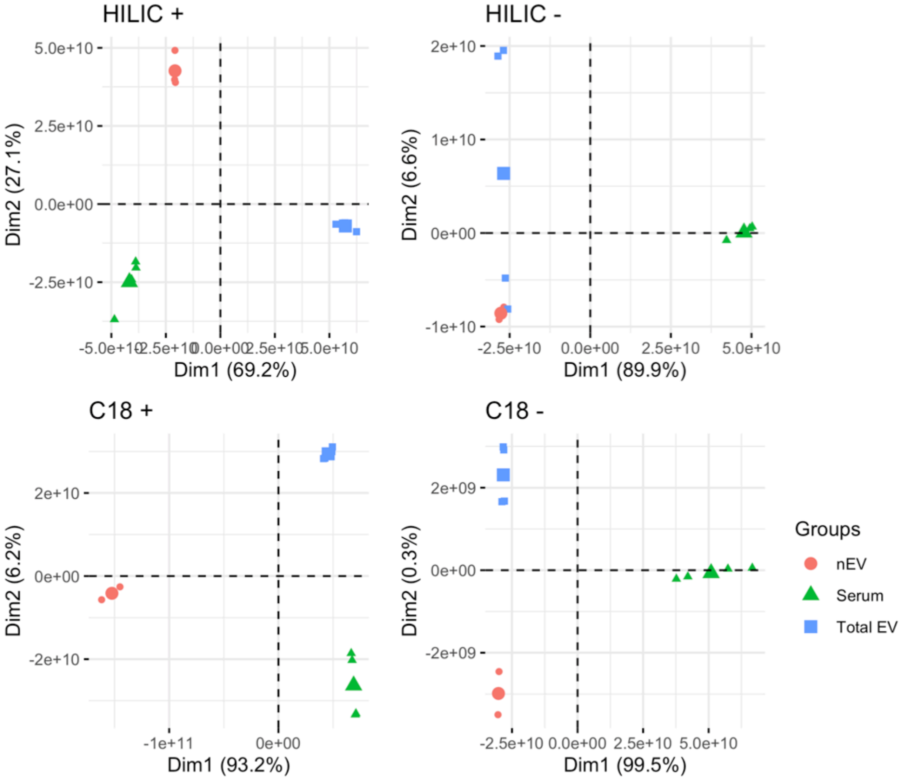

Supplemental Figure 4. Levels of each metabolite identified as being higher in nEVs compared to either total EVs or serum. The first label (on top) indicates the sub class and the second indicates the feature annotation.

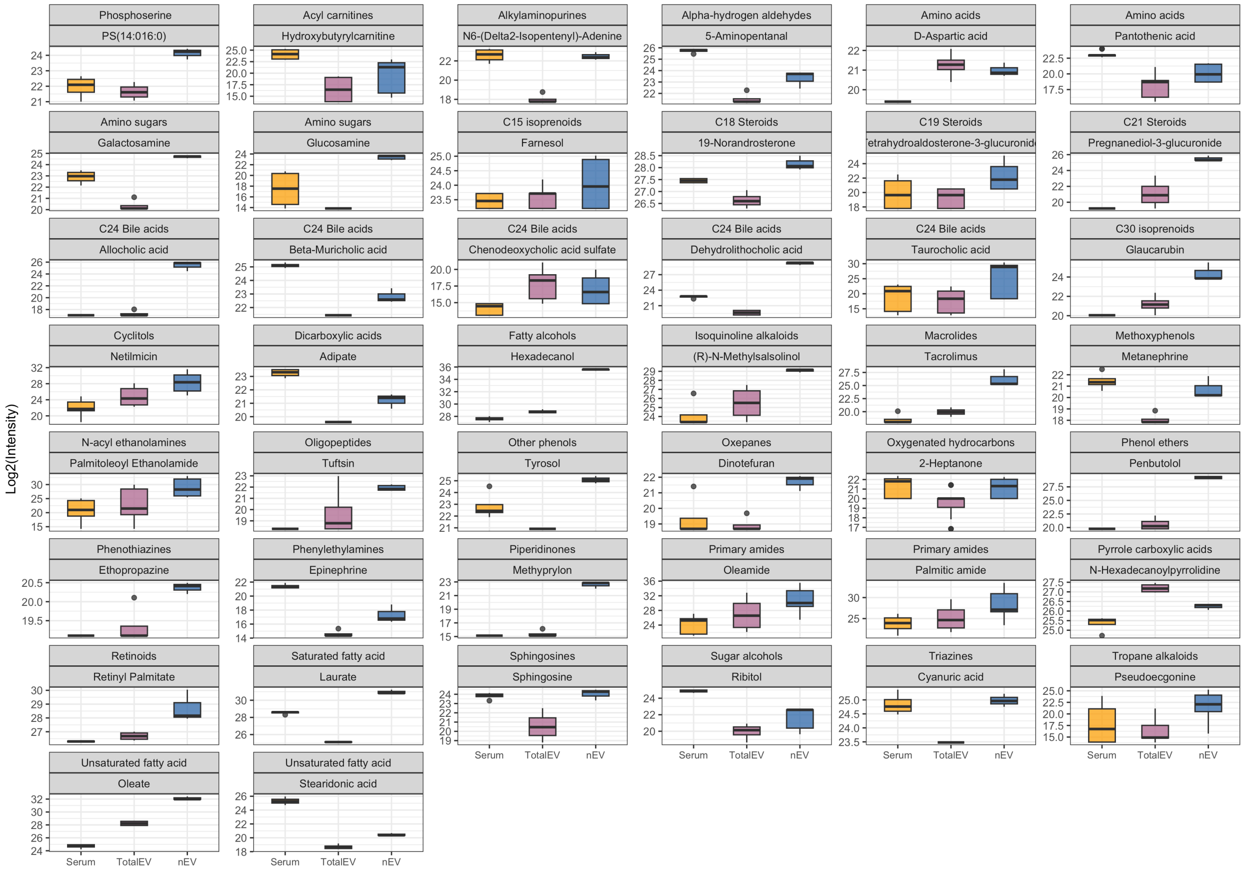
